# Population disruption: estimating changes in population distribution of the UK during the COVID-19 pandemic

**DOI:** 10.1101/2021.06.22.21259336

**Authors:** Hamish Gibbs, Naomi R Waterlow, James Cheshire, Leon Danon, Yang Liu, Chris Grundy, Adam J. Kucharski, LSHTM CMMID COVID-19 Working Group, Rosalind M. Eggo

**Affiliations:** Department of Infectious Disease Epidemiology, London School of Hygiene & Tropical Medicine, London, United Kingdom; Department of Geography, University College London, London, United Kingdom; Department of Engineering Mathematics, University of Bristol, Bristol, United Kingdom; The Alan Turing Institute, British Library, London, United Kingdom; Bristol Vaccine Centre, University of Bristol, Bristol, United Kingdom

## Abstract

Mobility data have demonstrated major changes in human movement patterns in response to COVID-19 and associated interventions in many countries. This can involve sub-national redistribution, short-term relocations as well as international migration. In this paper, we combine detailed location data from Facebook measuring the location of approximately 6 million daily active Facebook users in 5km^2^ tiles in the UK with census-derived population estimates to measure population mobility and redistribution. We provide time-varying population estimates and assess spatial population changes with respect to population density and four key reference dates in 2020 (First lockdown, End of term, Beginning of term, Christmas). We also show how population estimates derived from the distribution of Facebook users vary compared to mid-2020 small area population estimates by the UK national statistics agencies. We estimate that between March 2020 and March 2021, the total population of the UK declined and we identify important spatial variations in this population change, showing that low-density areas have experienced lower population decreases than urban areas. We estimate that, for the top 10% highest population tiles, the population has decreased by 6.6%. Further, we provide evidence that geographic redistributions of population within the UK coincide with dates of non-pharmaceutical interventions including lockdowns and movement restrictions, as well as seasonal patterns of migration around holiday dates. The methods used in this study reveal significant changes in population distribution at high spatial and temporal resolutions that have not previously been quantified by available demographic surveys in the UK. We found early indicators of potential longer-term changes in the population distribution of the UK although it is not clear if these changes may persist after the COVID-19 pandemic.

## Introduction

Responding to the outbreak of the COVID-19 pandemic has involved the widespread use of location data collected from mobile devices^1–3^. Location data aggregated from individual GPS locations and Call Detail Records have been used as a proxy for social contact in infectious disease models^4^, and have been used to measure adherence to relevant non-pharmaceutical interventions, such as domestic movement restrictions^5^. These data are typically aggregated to preserve user privacy by decreasing the spatial and/or temporal resolution of the data and applying censoring thresholds to aggregated metrics^1^. The availability of these data sources has increased as platforms including Facebook, Apple, Google, CityMapper, and mobile network providers like Vodafone and BT, have shared anonymised user data to aid the response to the pandemic^6–8^.

The most common use of mobile location data for responding to COVID-19 has been as a measure of travel and activity. This data is typically aggregated, anonymised and expressed as a normalised deviation from a baseline value for specific locations or pairs of locations^1,2,5,9–11^. Personally identifiable location data are also used for contact tracing and to monitor individuals’ adherence to isolation and quarantine^1,12^. In the early stages of the pandemic, mobile location data were combined with epidemiological data to estimate the spatial diffusion of SARS-CoV-2 transmission^13,14^. Research using mobile location data has demonstrated the large impact of COVID-19 and associated control measures such as movement restrictions and “stay-at-home” orders on patterns of human movement^5,10,14–16^. While there is strong evidence of disruptions in relative volumes of travel between locations, there is less clarity about the specific spatial and temporal changes in population distribution that have occurred during the pandemic.

The way that the distribution of population changes through time has important implications for public health responses, health and economic impact assessments of COVID-19, and the understanding of epidemic severity in the event of large redistributions of population. Population estimates are critical for epidemiological analysis, but research typically employs static estimates of population derived from annual or bi-annual census surveys and projections. While these static population estimates are accurate at the time of measurement, they do not reflect changes caused by major disruptions to patterns of movement and migration^17^. In the UK, there have been a limited number of surveys attempting to estimate the distribution of population during the COVID-19 pandemic, primarily focusing on specific locations like London, or specific groups such as foreign workers^18,19^.

In this paper, we focus on creating estimates of changes in the UK population distribution at a high spatial and temporal resolution to address existing gaps in our understanding of the population distribution in the UK across continuous time scales. We propose a method for generalising the location of Facebook users to the entire UK population, and provide population estimates that “fill in” missing data between census population estimates. We show large disruptions in the population distribution of the UK in the first year of the COVID- 19 pandemic, with population relocations coinciding with seasonal patterns of movement and the dates of public health interventions. We also observe an overall trend of decreased populations in urban areas. We compare time-varying population estimates derived from Facebook users to mid-2020 population estimates from UK national statistics agencies, showing how within-year dynamic populations differ from annual population estimates. This has important implications for modelling approaches that employ static population estimates, as estimates may not reflect the time-varying nature of underlying populations.

## Methods

Facebook, a social network and mobile application provider, records users actively sharing their location with Facebook applications, referencing these locations to an approximately 2.5km^2^ gridded tile system in sequential 8-hour windows between March 10th 2020 and March 31st 2021^6^. We estimated changes in the UK population by: 1) aggregating tiles to a 5km^2^ grid to align with census population estimates; 2) using 2019 mid-year population estimates for small statistical areas (Supplemental Figure 1) to estimate the spatial distribution of the UK population before the epidemic using changes in the distribution of Facebook users to adjust these population estimates each day^6,20^. The generalisation method assumes that Facebook users travel at the same rates as the rest of the UK population.

We measured spatial changes in population, focusing on four important dates in 2020: 1) the first national lockdown introduced on March 23rd, 2020; 2) the beginning of summer holidays on July 21st, 2020; 3) the return to school on September 1st, 2020; and 4) Christmas on December 25th, 2020. School start and finish dates are different in the nations of the UK and regionally, so we selected approximate values for schools in England. Note that schools in the UK were not open for in-person learning between March and July 2020 due to COVID-19 restrictions.

### Population Data

We used population data provided by the Facebook Data for Good program^6^ which records the number of active Facebook users in spatial tiles in sequential time periods. We used data from March 10th 2020 to March 31st 2021, and we also rely on the Facebook-generated baseline from a 45 day period between January 29th and March 9th 2020 as our reference for the baseline population of Facebook users.

Facebook datasets are provided in a gridded spatial reference, referred to as “tiles” which are referenced by a unique integer (a “quadkey”). The resolution of tiles is defined by a “zoom level.” The population data is provided at zoom level 13 (approximately 2.5 km^2^) and zoom level 12 (approximately 5 km^2^). We aggregate population data to zoom level 12 to combine it with census population estimates. There are 4 level 13 tiles for each level 12 tile. Data are provided in sequential 8-hour periods (00:00 - 08:00, 08:00 - 16:00, and 16:00 to 00:00) and are available with approximately 48 hours delay. Baseline population values are defined as the median number of users in a tile for each 8-hour period during a baseline period. These values provide an aggregated measure of the population of individual tiles, which we use to incorporate information preceding the start of the full dataset on March 10th 2020.

We applied a population adjustment to the population of Facebook users, transforming the number of Facebook users into the number of actual people using 2019 mid-year population estimates from small statistical areas in the UK (England and Wales: Output Areas, Northern Ireland: Small Areas, Scotland: Data Zones). These estimates are the highest resolution population data produced by UK statistics agencies. We used 2019 mid-year estimates as they are the closest estimates available to the baseline period of the Facebook data. There is uncertainty in the relationship between 2019 population estimates and the distribution of Facebook users due to unobserved changes in population between 2019 and March 2020.

Facebook population data do not include demographic information and therefore, we are not able to compare to the total UK population on age structure, gender, ethnicity, or socioeconomic status because of the privacy-preserving structure of the dataset. In previous research, we have found no associations with key demographic factors at the tile level^24^. Users may also be removed from the dataset due to varying patterns of usage including deactivating their accounts or pausing their use of Facebook services, or because of international travel. Finally, because location sharing is an “opt-out” feature for Facebook users, there may be bias in the population of Facebook users included in these datasets. We use data from a specific time period (16:00 to 00:00), the period with the lowest weekly variance in the total number of Facebook users included in the data set (Supplemental Figure 2).

### Population Adjustment

We estimated the UK population from the number of Facebook users by combining Facebook population data with census population estimates in Bing tiles, a regular spatial grid of approximately 4.8km^2^. First, we extracted census Population estimates to grid cells by assigning the population weighted centroid of each census to the overlapping tile. We then summed the census population estimates for each tile

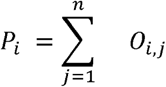

where *p*_*i*_ is the census population of the *i*^*th*^ tile, and,, is the *j*^*th*^ OA population estimate intersecting the *i*^*th*^ tile. We then computed the the median number of Facebook users in each tile in the baseline period for each tile

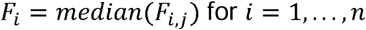

where *F*_*i*_ is the reference number of Facebook users in the *i*^*th*^ tile and *F*_*i,b*_, is the baseline value for the the *i*^*th*^ tile. We then computed the proportion of Facebook users to census population for each tile

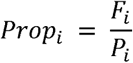

where *prop*_*i*_ the is the proportion of Facebook users, *F*_*i*_ is the baseline number of Facebook users, and *p*_*i*_ is the census population estimate of the *i*^*th*^ tile. We then adjusted the number of Facebook users to the population using these proportions

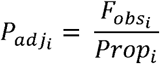

where 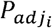 is the adjusted population, 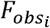 is the observed number of Facebook users, and *prop*_*i*_ is the baseline proportion of Facebook users to census population for the *i*^*th*^ tile.

### Population Variation

We measured the change in population in individual cells in sequential daily and weekly time periods as a proxy for population movement. To compute this measurement in daily periods, we computed the difference between lagged and current population estimates for individual tiles. We then summed the absolute value of these differences for daily periods.

To compute the difference in population change for weekly periods, we computed the average population of individual tiles per week, then calculated the difference between lagged and current population estimates for individual tiles. We summed the absolute value of these differences for weekly periods to measure weekly changes in population.

We assigned tiles to population deciles based on the value of their census populations. This labelling was consistent across the time series and was not altered in response to population changes in individual tiles.

### Mid-year Census Comparison

To determine whether generalised population estimates accurately reflected the population distribution of the UK as measured by the census, we compared these generalised estimated to mid-2020 population estimates from UK national statistics agencies. Mid-year population estimates describe the resident population on a specific date: 30th June 2020. We compared estimates on this date in individual grid cells and in population deciles. Population deciles were defined by mid-2019 population estimates.

### COVID-19 Incidence Rates

To compare how COVID-19 incidence rates are affected by the underlying population estimates, we aggregated our Facebook-derived population estimates to the same spatial scale as reported COVID-19 data. We used publicly-available data on confirmed COVID-19 cases by specimen date in Lower Tier Local Authorities (LTLAs)^27^. We aggregated this data to Built Up Areas (BUAs) using a lookup of LTLAs to BUAs^28^. We restricted this method to the top 20 BUAs by population (total BUAs: 6,347) because these BUAs encompassed multiple LTLAs, ensuring spatial consistency between COVID-19 case data and population data.

We then extracted census and dynamic (estimated) population data from tiles to BUAs. We computed the rate of COVID-19 cases per 100k people against a static population for each time period using

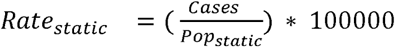

Where *Rate*_*static*_ is the rate of COVID-19 cases *Cases* in a given time period against a static population *Pop*_*static*_ We computed the same rate using a time-varying (dynamic) population using:

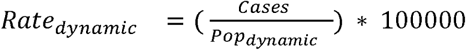

Where *Rate*_*dynamic*_ is the rate of COVID-19 cases *Cases* in a given time window by population, *Pop*_*static*_.

## Results

### Population Data

To understand the usual spatial distribution of Facebook users, we used Facebook-generated baseline estimates of population calculated in the 45 days before March 10th, from January 25th to March 9th, 2020. This baseline period is automatically back-calculated by Facebook prior to data sharing and provides the earliest available information on the distribution of Facebook users in the dataset. Before applying the population adjustment, we compared the population of Facebook users in the baseline period to census population estimates across the UK to identify spatial variations in the distribution of Facebook users and census population (Figure 1a, Supplemental Figure 1). We found a different median percentage of Facebook users to population in each of the three 8-hour reporting periods (00:00 to 08:00, 08:00 to 16:00, and 16:00 to 0:00), of 7.46%, 9.1%, and 9.77% respectively (Figure 1b, c). These distributions reflect differences in the number of people actively using Facebook services at different times of day. During the baseline period, we found that the majority of tiles (77%) have percentages of Facebook usage between 5 and 20% (Supplementary Figure 3). There are some tiles where Facebook usage exceeds 75%, which was typically observed in low population tiles. Extreme discrepancies greater than 100% Facebook usage, observed in 33 tiles in total, may result from interference from sparse cellular network coverage (Supplemental Figure 4).

**Figure 1.**
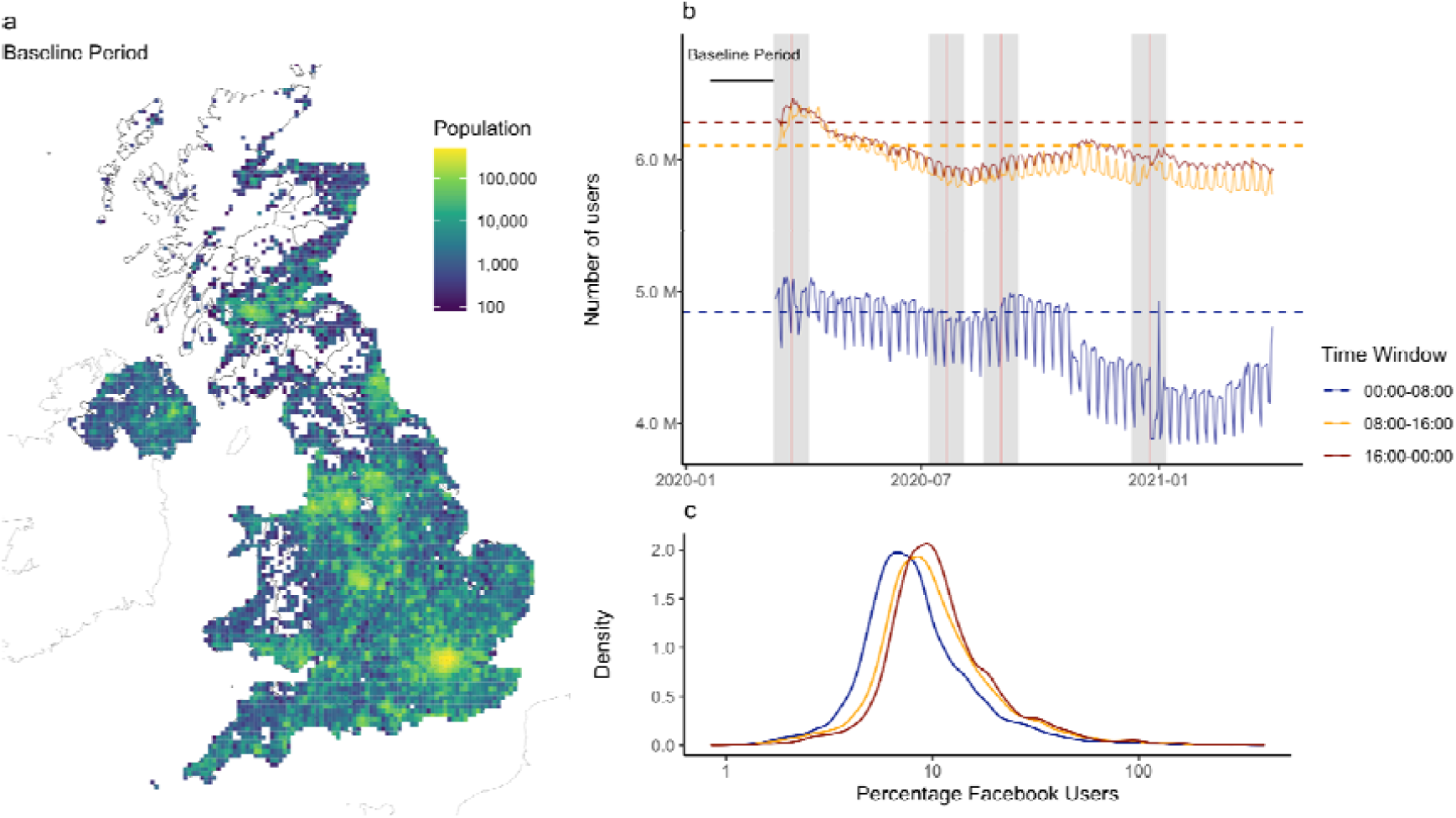
The relationship between Facebook users and census population estimates. a) The population of the UK in the baseline period estimated by generalising the locations of Facebook users actively sharing location. b) The total number of Facebook users in each time window, dashed horizontal lines show baseline values used to define the proportion of users to census population. Note that axis limits for the number of users do not begin at 0. Grey bars indicate the 2 weeks preceding and following reference dates (red lines): “First national lockdown”, “beginning of summer holidays”, “return to school”, and “Christmas”. The decrease in users in the 00:00 to 08:00 period in late October most likely results from daylight savings time, which is not accounted for in these time windows. c) The distribution of the percentage of Facebook users in individual cells for each daily time window, showing that Facebook usage as a proportion of population tends to be higher in daytime periods.

### Dynamic population changes

We observed decreases in the population of urban areas following the announcement of the first national lockdown, with a corresponding increase in less densely populated areas (Figure 2a, Supplemental Figure 5). This pattern is similar to the population changes observed in the beginning of the summer holidays period, when populations decreased in populous urban and suburban areas (Figure 2b). We observed an inverse pattern of population change during the return to school period, where populations increased in populous and decreased in less populous areas (Figure 2c). The Christmas period coincided with an announcement of travel restrictions in the South East of England on December 19th, 2020 (Tier 4), with a short term relaxation of measures on Christmas day for other areas^21^ (Supplemental Figure 6). In this period, we observed a large decrease in population in central London and an increase in the population in tiles immediately peripheral to London (Figure 3d).

**Figure 2.**
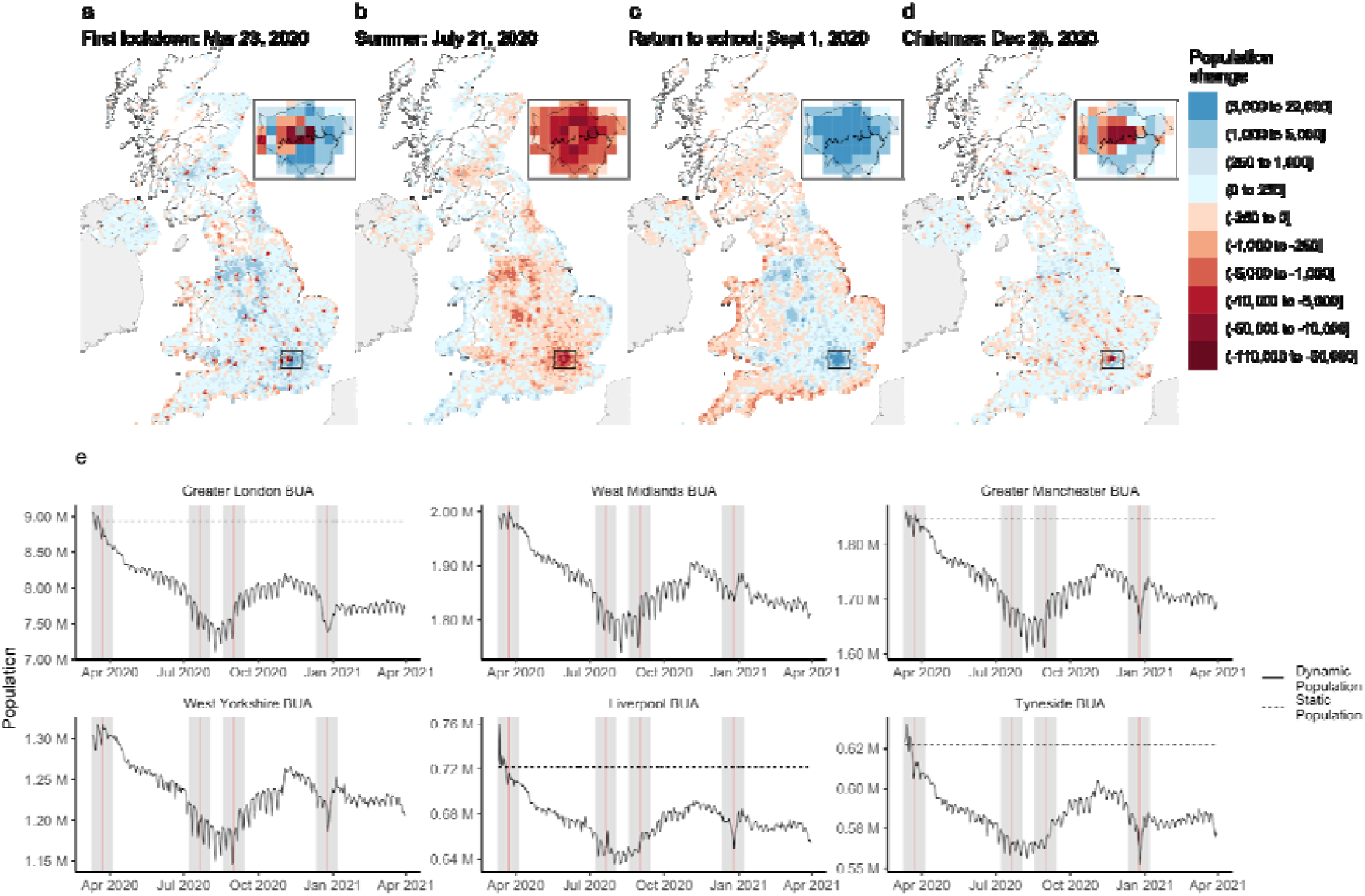
Time-varying estimates of population change. Population change in the two weeks preceding and following significant reference dates: a) “First national lockdown”, b) “Summer”, c) “Return to school”, and d) “Christmas”. e) Time-varying population estimates for the six largest BUAs. Red lines indicate reference dates and grey areas show the two weeks preceding and following these dates.

**Figure 3.**
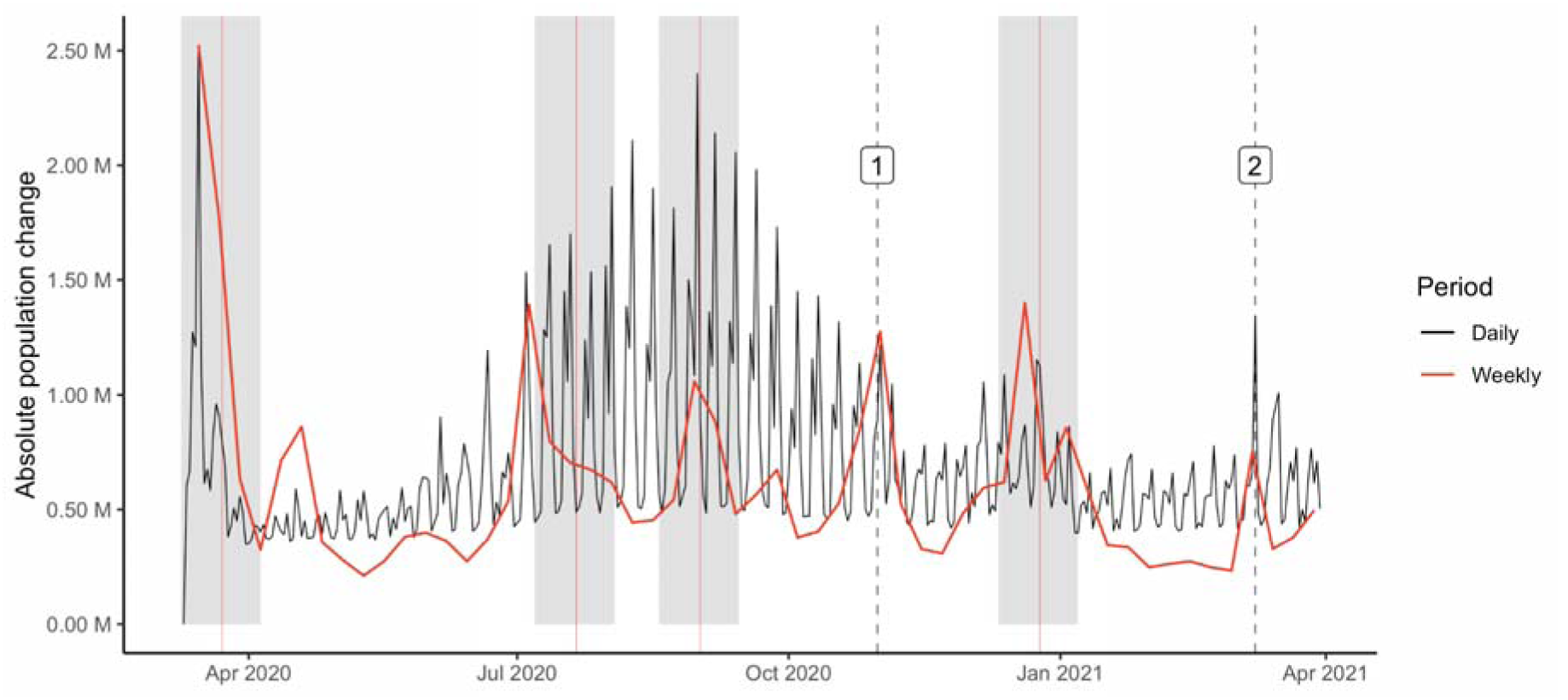
Total displacement of population through time. The total change in population through time, measured as the difference between lagged population estimates in daily and weekly time windows. Vertical dashed lines show significant dates ([1] Second National Lockdown, [2] Reopening of schools). Grey shading indicates the two weeks preceding and following these dates.

We calculated time-varying population estimates for the largest built-up areas (BUAs) in the UK which show major changes at key points in the epidemic (Figure 2e, Supplementary Figure 7-8). We observed a sustained decrease in the population of London between March 10th, 2020 and the implementation of public health measures on March 23rd, 2020. Comparing the 14 days before and after the first national lockdown, the population of London decreased by 3.44% (196,000 people). Further, we estimate the population of London was 646,000 lower in total between March 2020, and March 2021, with some periods of even lower population observed during the summer and Christmas holidays. Between March 2020 and March 2021, the population decreased in the 20 largest BUAs in the UK, with the largest decreases observed in Greater London, West Midlands, and Greater Manchester BUAs.

The absolute daily difference in the population size of each tile indicates the daily degree of population flux. The total absolute daily difference across the UK had strong short-term periodicity, but on a weekly level, changes in flux were clearly visible in association with the four key reference dates (Figure 3). Additionally, other national interventions also showed responses with this metric, including the second national lockdown on November 2nd 2020 marked with a 1, and the reopening of schools on March 8th (schools in the UK were shut after January 1st 2020) marked with a 2.

### Rural population increase

We measured population changes in areas of different population density by dividing tiles into deciles based on their census population size (Supplemental Figure 8) and calculating time-varying population estimates for these deciles (Figure 4a). We found large decreases in the most populous decile in the first lockdown period, with the greatest increase in population observed in the 2nd most populous decile (Figure 4b). In the summer, we observed a decrease in the top two most populous deciles and a more even distribution of population among the remaining deciles, including the least populous. This reflects movement to rural areas coinciding with the beginning of summer holidays. The summer pattern is mirrored by a decrease in less populous tiles in the return to school period. Finally, we observed a similar, though reduced pattern of population decrease in the Christmas period where population in the most populous decile decreased, while in less populous deciles it increased.

**Figure 4.**
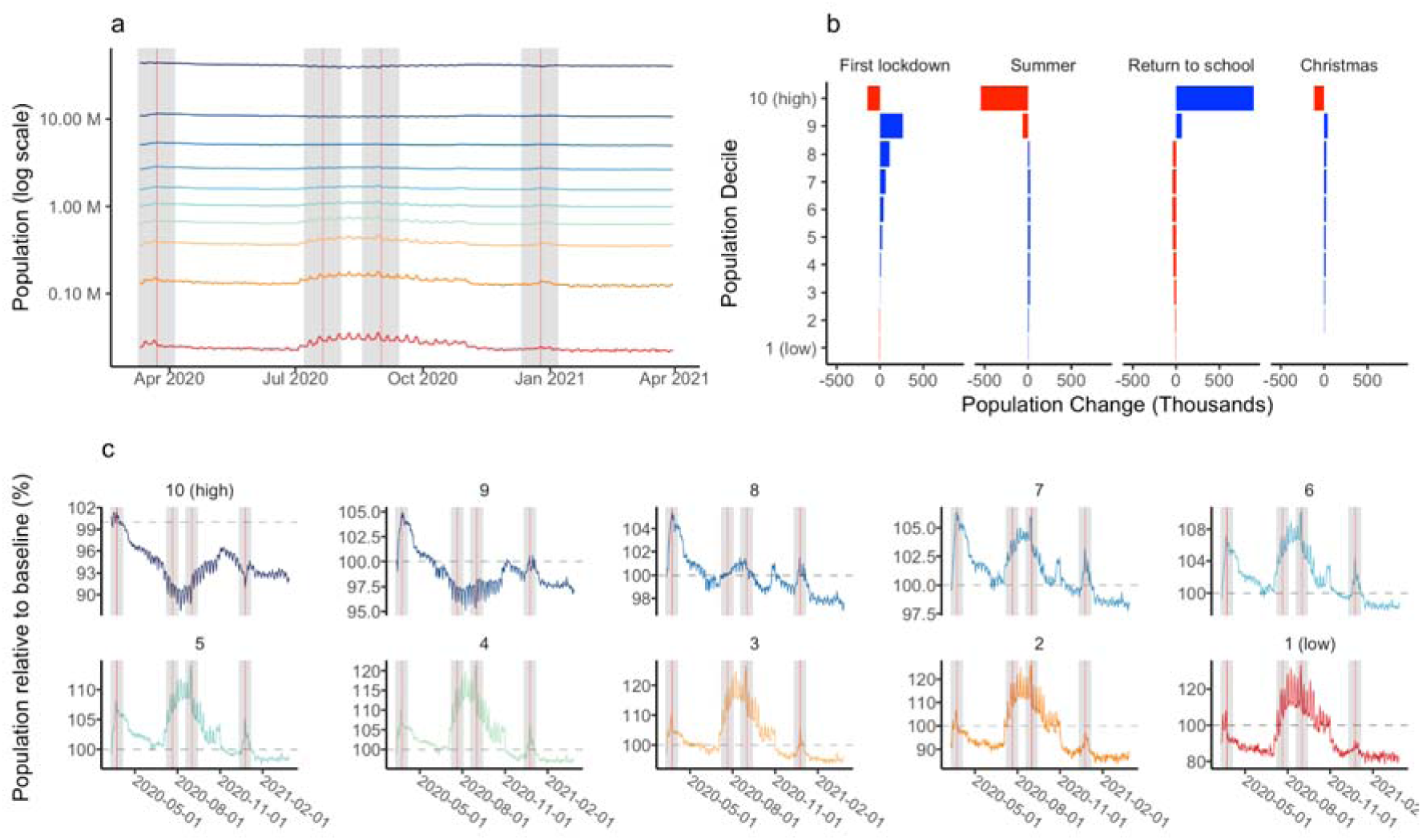
Population changes by population decile. a) The total population of tiles in each decile over time (log scale). b) The change in population for tiles in each population decile in the two weeks preceding and following reference dates. c) Population change in deciles relative to baseline population estimates. Grey shading indicates the two weeks preceding and following these dates. Note different y-axes in each plot.

Each decile showed different patterns of population change over the course of the study period (Figure 4c), where the lowest decile’s population increased to 120% of census estimates during the summer period. This decile shows a smaller response to Christmas than others, with the most pronounced increases in the central deciles. Of note is that most deciles did not return to the pre-Christmas population sizes, suggesting that people could have remained where they travelled for Christmas as the UK entered new restrictions on 26th December. We estimated that, for the top 10% highest population tiles, the population has decreased by 6.6%.

### Comparison to mid-2020 census estimates

We compared population estimates derived from the locations of Facebook users to 2020 mid-year population estimates from UK national statistics agencies (Figure 5a). The distribution of Facebook users estimated a higher proportion of the population outside of urban areas compared to mid-year 2020 census estimates. Because the Facebook population was generalized from mid-2019 census population estimates, the total population of the UK was underestimated compared to mid-2020 census population estimates by 2,899,000 individuals (4%), relating to an overall decline in the population of Facebook users. However, Facebook estimated a reduction of 2,635,000 (6%) in the highest population decile compared to mid-2020 census estimates which may indicate a redistribution of UK population out of the populous areas which is not captured in census estimates.

**Figure 5.**
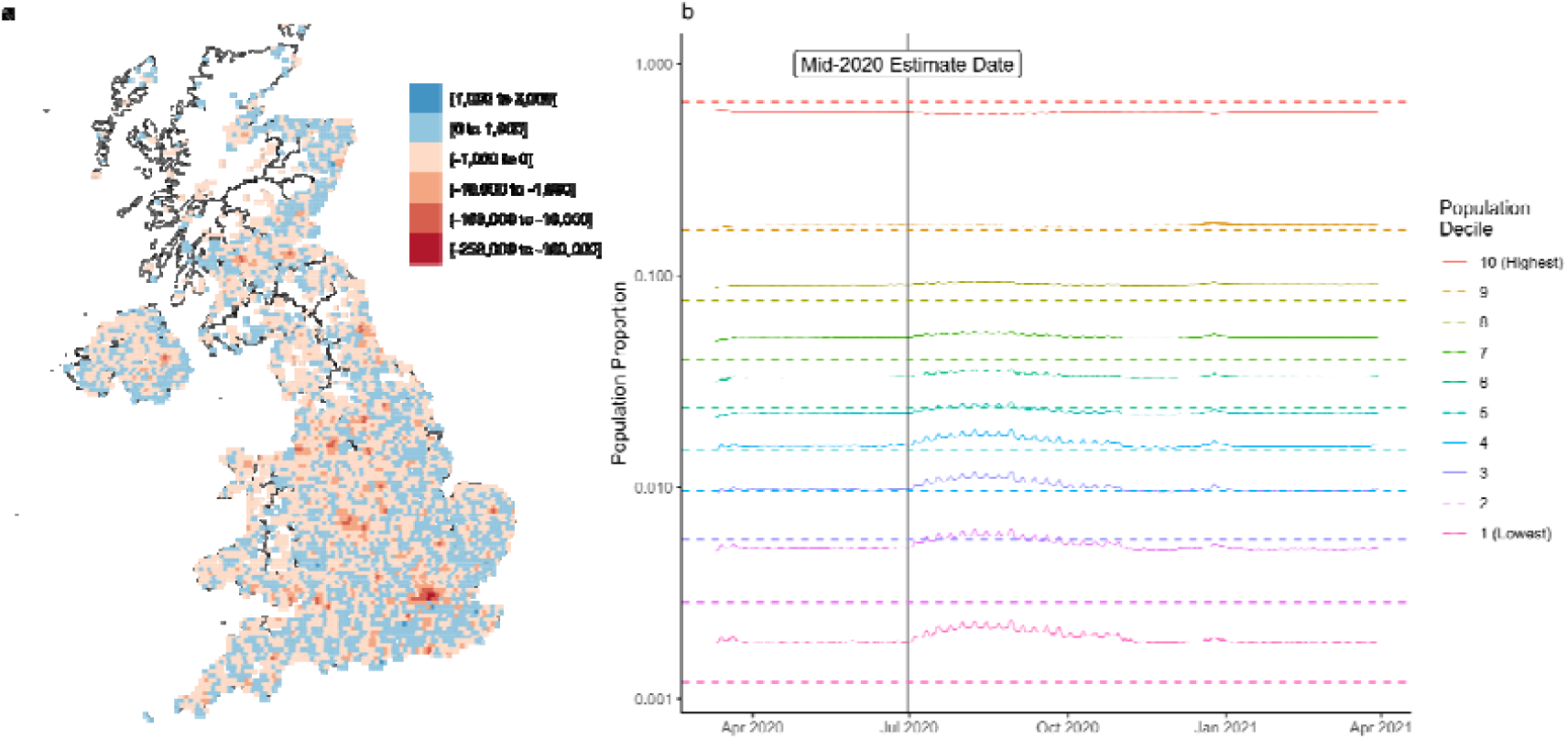
Comparison of generalized Facebook population to 2020 mid-year population estimates. a) Population difference between Facebook generalized population estimates and 2020 mid-year census estimates in tiles. Facebook data shows higher populations outside of urban areas than estimated in 2020. b) The proportion of Facebook estimated population in population deciles compared to 2020 population estimates. Dashed lines show 2020 mid-year population estimates. Black vertical line shows the date of 2020 mid-year population estimates.

### Effect on COVID-19 incidence estimates

Deviations between dynamic and census populations can also affect estimates of the impact of the epidemic in real-time. Some statistics, like the rate of cases per 100,000 rely on census population estimates to calculate disease incidence relative to a population. We found that the inclusion of time-varying population estimates led to changes in the calculated COVID-19 incidence rates, leading to an altered interpretation of the severity of outbreaks in different areas (Figure 6). Using time-varying population estimates, we identified differences in the rate of confirmed COVID-19 cases in the 8 largest BUAs in England ranging between 23.24% and −3.26% of the rate calculated with static populations (Supplemental Figures 12- 13). This difference between epidemic rates reflects changes in the underlying population of these areas through time. 84% of the time-varying rates are within 10% of the rates computed using a static population. Nonetheless, the differences in epidemic rates are not trivial, particularly in urban areas during the period of highest COVID-19 cases between December 2020 and January 2021. The combined effect of high numbers of cases and population outflows from urban areas led to a consistent underestimation of the scale of the COVID-19 epidemics in the most populous Built-Up Areas (Supplemental Figures 10-11).

**Figure 6.**
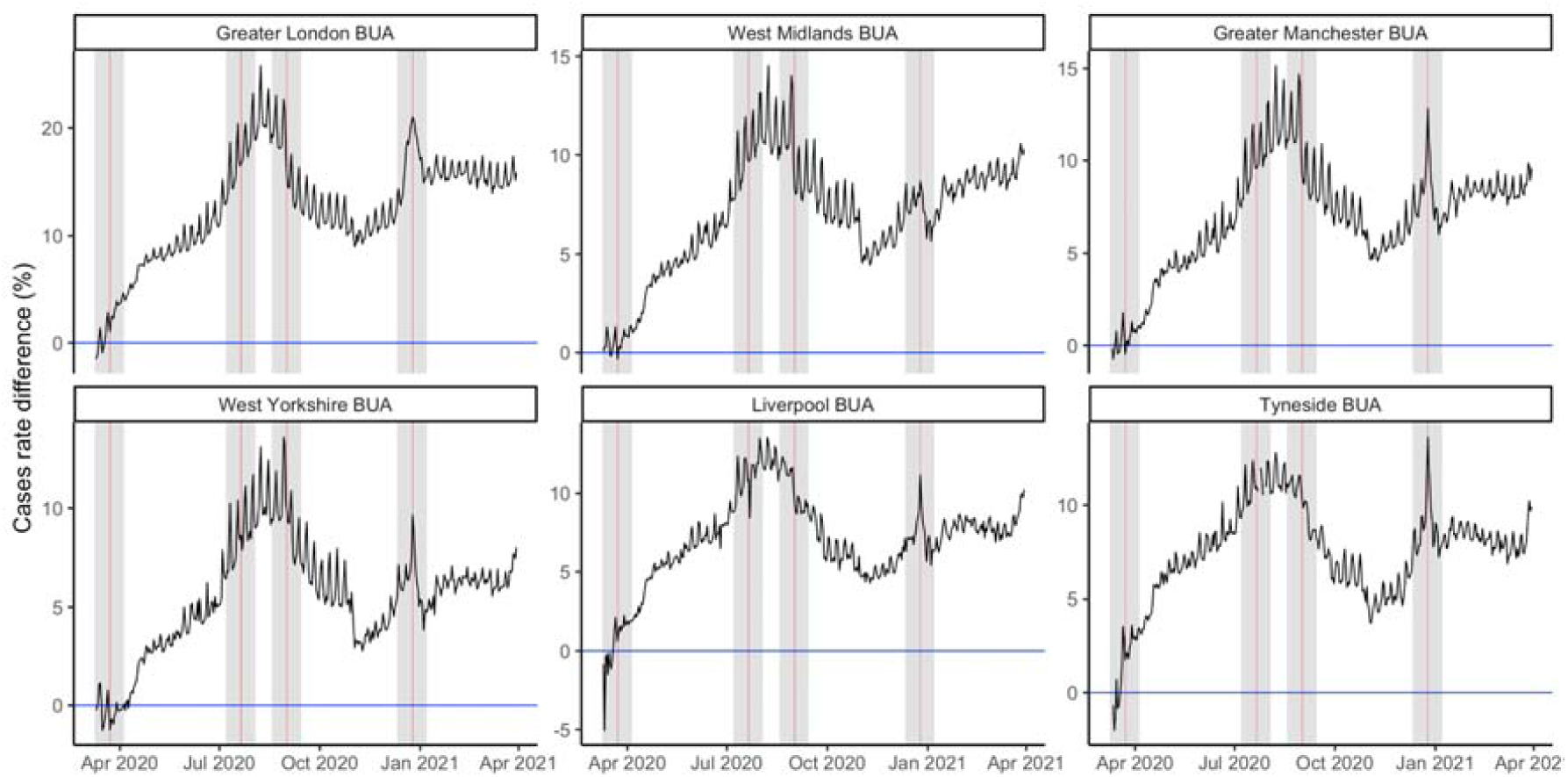
Difference in case rates by population estimation. The percentage difference between the rate of COVID-19 cases per 100,000 in Built Up Areas calculated using static census populations (blue) and dynamic populations (black). Vertical red lines show reference dates. Grey shading indicates the two weeks preceding and following these dates

## Discussion

In this study, we present a novel approach for estimating population changes using a large, near-real time dataset of the location of millions of Facebook users in the UK. This study identifies important changes in the population of the UK coinciding with the announcement of public health interventions and with seasonal migrations in the UK. We provide evidence supporting previous reports of a decrease in population in urban areas. We also demonstrate how observed changes in population have varied in space and time. We show how these population changes can impact disease transmission and the predicted size of epidemics. This is particularly important for understanding how the timing of population changes relate to the progression of a disease epidemic. We also provide new estimates of the rate of cases using time-varying populations in different locations, which could be extended to other rates like calculated rates of vaccinations.

The COVID-19 pandemic has resulted in unprecedented changes in the population distribution of the UK. These changes include abrupt changes in population as well as broad trends of population change since the announcement of the first national lockdown in March 2020. Decreases in the population of the 20 largest urban areas in the UK have persisted throughout 2020 which may reflect new patterns of employment and home working. The trend of decreasing urban populations has not been monotonic, as the population of urban centers has increased during specific periods. Nor has the trend of population increases in less populous areas been uniform. As we demonstrate, rural populations have experienced unique population dynamics during public health interventions and seasonal migrations during summer holidays.

Our estimates of population change are similar to the limited available estimates of population changes during COVID-19 in the UK, but provide much higher spatial and temporal resolution^18,19^. In January 2021, the consulting firm PWC estimated that the population of London would decrease by 300,000 people, while a study of the non-UK born population estimated a decrease of ∼600,000 people in Greater London^18,19^. While these estimates are limited, they provide helpful early evidence of the large-scale changes in population that have occurred during the pandemic and reflect estimated population changes of a similar magnitude as those reported in detail in this study (646,000 in Greater London).

The population estimates calculated in this study are a novel resource for understanding the evolution of the UK population through time and could be used to “fill the gap” between census population estimates. In the future, these estimates can be compared to results from the 2021 census to gain a detailed understanding of their accuracy at a high spatial resolution, and to identify potential biases in the generalization method. It is important to note that demographic trends are informed by varying rates of births, deaths, immigration and emigration. Changes in commuting patterns, the labor market, and seasonal migrations also impact patterns of population change^17,19^. The estimates presented in this study are intended to provide detailed information on dynamic population changes between March 2020 and March 2021 but it is unclear how the trends of population change observed during the COVID-19 pandemic will persist in the future.

We demonstrated that time-varying population estimates impact reported rates of disease incidence. Other measurements of disease impact (attack rate, hospitalisation rate) could also be calculated by incorporating contemporaneous (time-varying) estimates of population. While this information is useful for retrospective reinterpretations of the severity of disease in particular locations, it would be particularly valuable for the modelling of disease spread, where time-varying population estimates could more accurately reflect population distributions in real time.

This study and the proposed method of population generalisation have a number of limitations. As the basis for our population generalisation, we use the closest available census population estimates to the baseline period. As demonstrated in this study, the population of the UK experiences dynamic redistributions and it is not possible to identify any population changes which occurred between the time of census estimation and the baseline period. The estimates presented in this study will be valuable for comparison to results from the 2021 census and can provide further information on the use of alternative sources of population data for measuring patterns of population change.

Further research is required to fully understand the demographic characteristics of Facebook users who are presented in aggregated population and mobility metrics, and how the behaviour of these individuals varies from the general population^22,23^. There is still a limited understanding of how user subsets from applications like Facebook vary from the general population and how this difference may be reflected in aggregated location metrics. In the future, research on the bias of these user subsets could be used to improve the generalisation of the behaviour of these individuals for representing the entire population.

## Conclusion

Time-varying population estimates provide detailed information on major changes in the population distribution of the UK during the COVID-19 pandemic. Generalising the movement of Facebook users, we present strong evidence of population decreases in major urban areas in the UK.

## Data Availability

The terms of use of data from the Facebook Data for Good program prohibit unauthorised distribution. Data is available from the Facebook Data for Good Partner Program by application. Data processing and analysis code is available from https://github.com/hamishgibbs/facebook_population_2020_2021. We have also aggregated our population estimates to 2019 Local Authority Districts, available from https://zenodo.org/record/5013620.

https://zenodo.org/record/5013620

## Data Availability

The terms of use of data from the Facebook Data for Good program prohibit unauthorised distribution. Data is available from the Facebook Data for Good Partner Program by application. We have aggregated our population estimates to 2019 Local Authority Districts, available from https://zenodo.org/record/5013620.

## Code Availability

Data processing and analysis code is available from https://github.com/hamishgibbs/facebook_population_2020_2021.

## Acknowledgements

The following authors were part of the Centre for Mathematical Modelling of Infectious Disease COVID-19 Working Group. Each contributed in processing, cleaning and interpretation of data, interpreted findings, contributed to the manuscript, and approved the work for publication: Damien C Tully, Alicia Rosello, David Hodgson, Kaja Abbas, Gwenan M Knight, Sam Abbott, Katherine E. Atkins, Christopher I Jarvis, Kiesha Prem, Matthew Quaife, Graham Medley, Stefan Flasche, Frank G Sandmann, Rosanna C Barnard, William Waites, Nikos I Bosse, Rachel Lowe, C Julian Villabona-Arenas, Simon R Procter, W John Edmunds, Mark Jit, Ciara V McCarthy, Mihaly Koltai, Akira Endo, Billy J Quilty, Rachael Pung, Oliver Brady, Amy Gimma, Timothy W Russell, Carl A B Pearson, Stéphane Hué, Fiona Yueqian Sun, Joel Hellewell, James D Munday, Emilie Finch, Samuel Clifford, Katharine Sherratt, Paul Mee, Nicholas G. Davies, Lloyd A C Chapman, Kathleen O’Reilly, Sophie R Meakin, Yalda Jafari, Kerry LM Wong, Sebastian Funk.

