## Supplementary Information for "Population disruption: estimating changes in population distribution of the UK during the COVID-19 pandemic"

**Affiliations:**

^†^ Membership of LSHTM CMMID COVID-19 working group is provided in the acknowledgments.

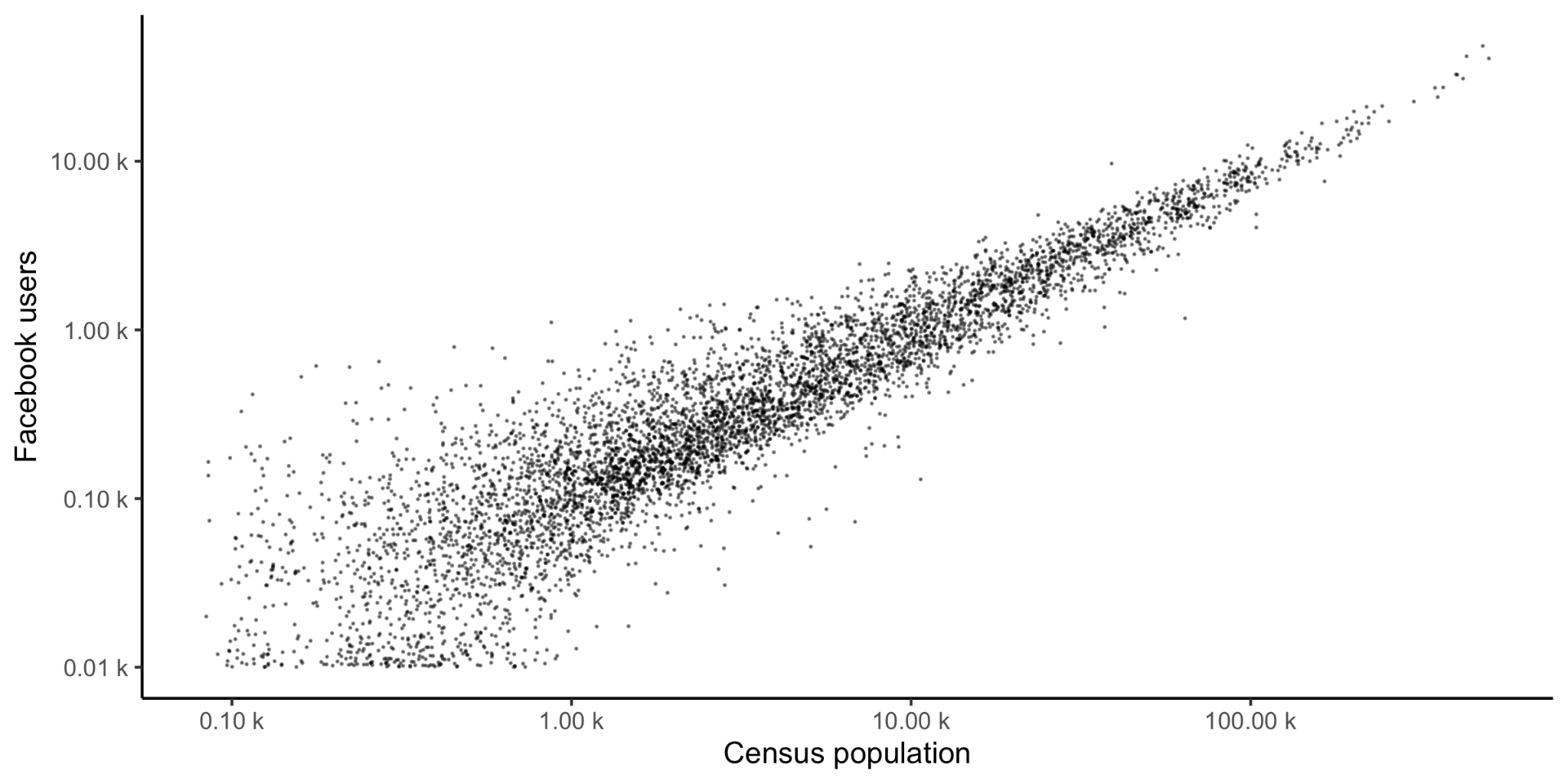

**Supplemental Figure 1. Comparison of Facebook users and census population.** The population of Facebook users in the 16:00 to 00:00 baseline period and census population estimates in individual tiles (log_10_ - log_10_ scale).

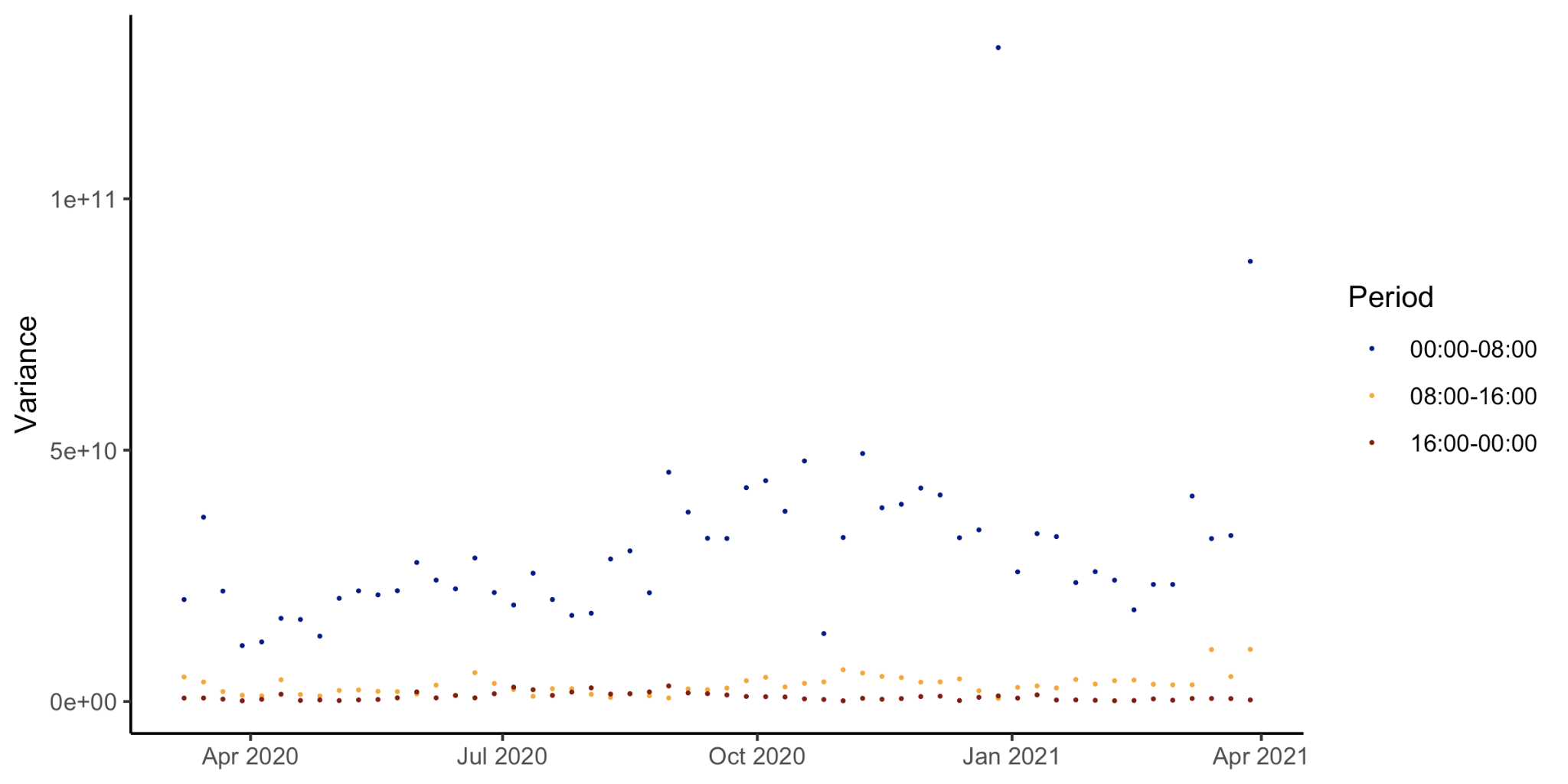

**Supplemental Figure 2. Weekly variance of the observed number of Facebook users.** The variance of the number of Facebook users in different time periods. The 16:00 to 00:00 period displays the lowest variance.

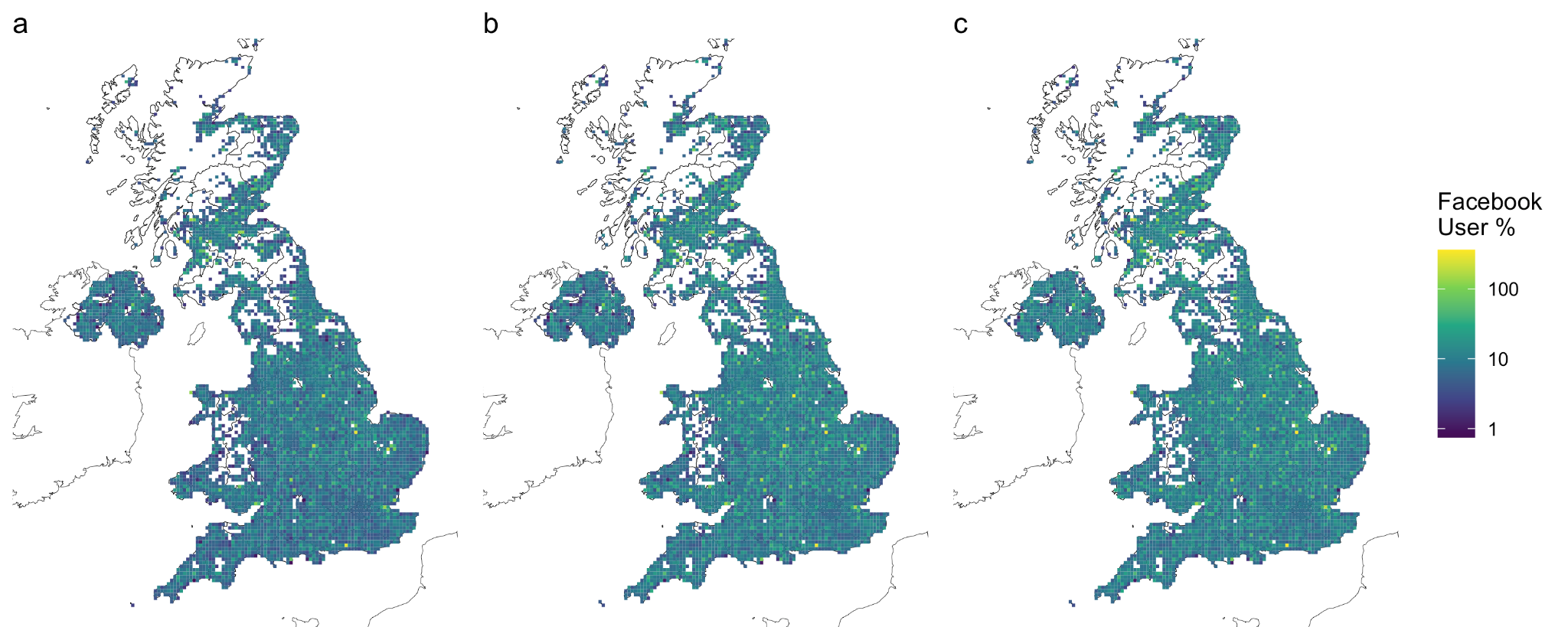

**Supplemental Figure 3. The percentage of Facebook users.** The percentage of Facebook users to Census population in individual tiles in each time window in the baseline period: a) 00:00 to 08:00, b) 08:00 to 16:00, c) 16:00 to 00:00.

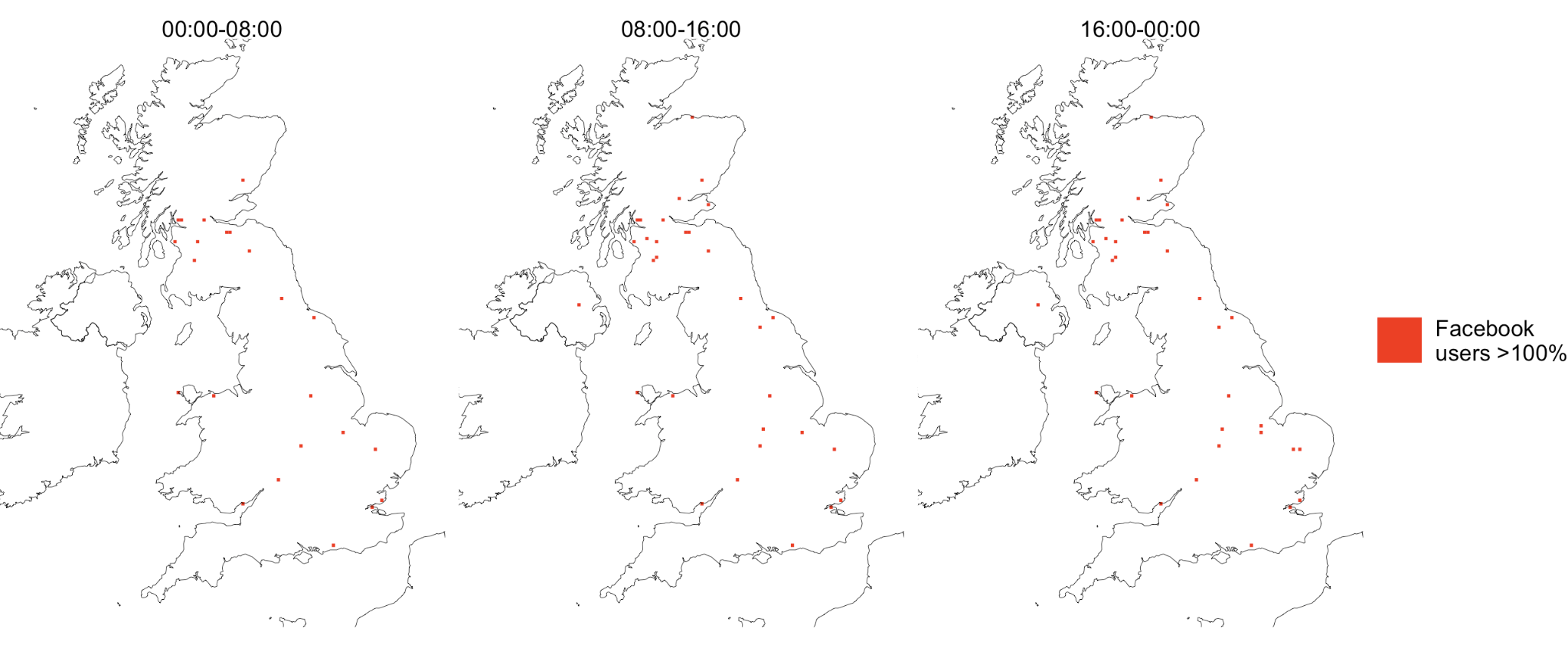

**Supplemental Figure 4. Cells with Greater than 100% Facebook Usage.** These cells are found in low population areas and may result from interference from sparse cellular network coverage.

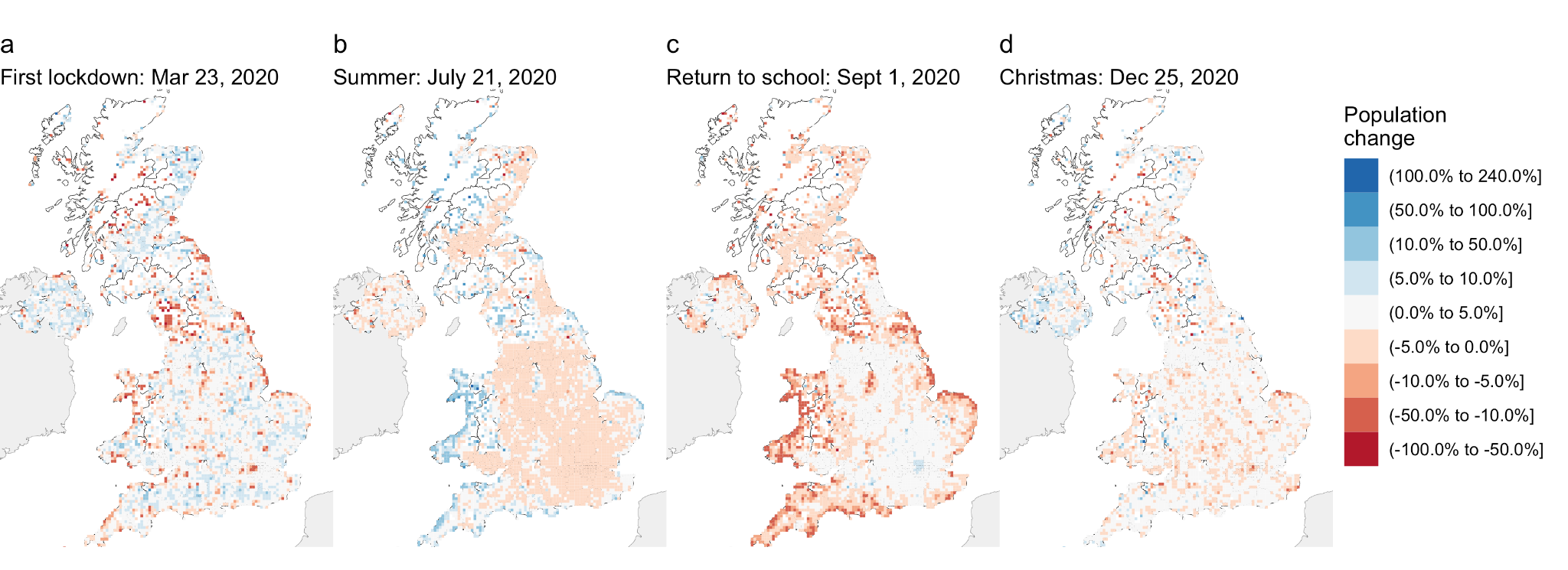

**Supplemental Figure 5. Time-varying estimates of % population change.** Percentage population change in the two weeks preceding and following significant reference dates: a) First national lockdown, b) Summer, c) Return to school, and d) Christmas.

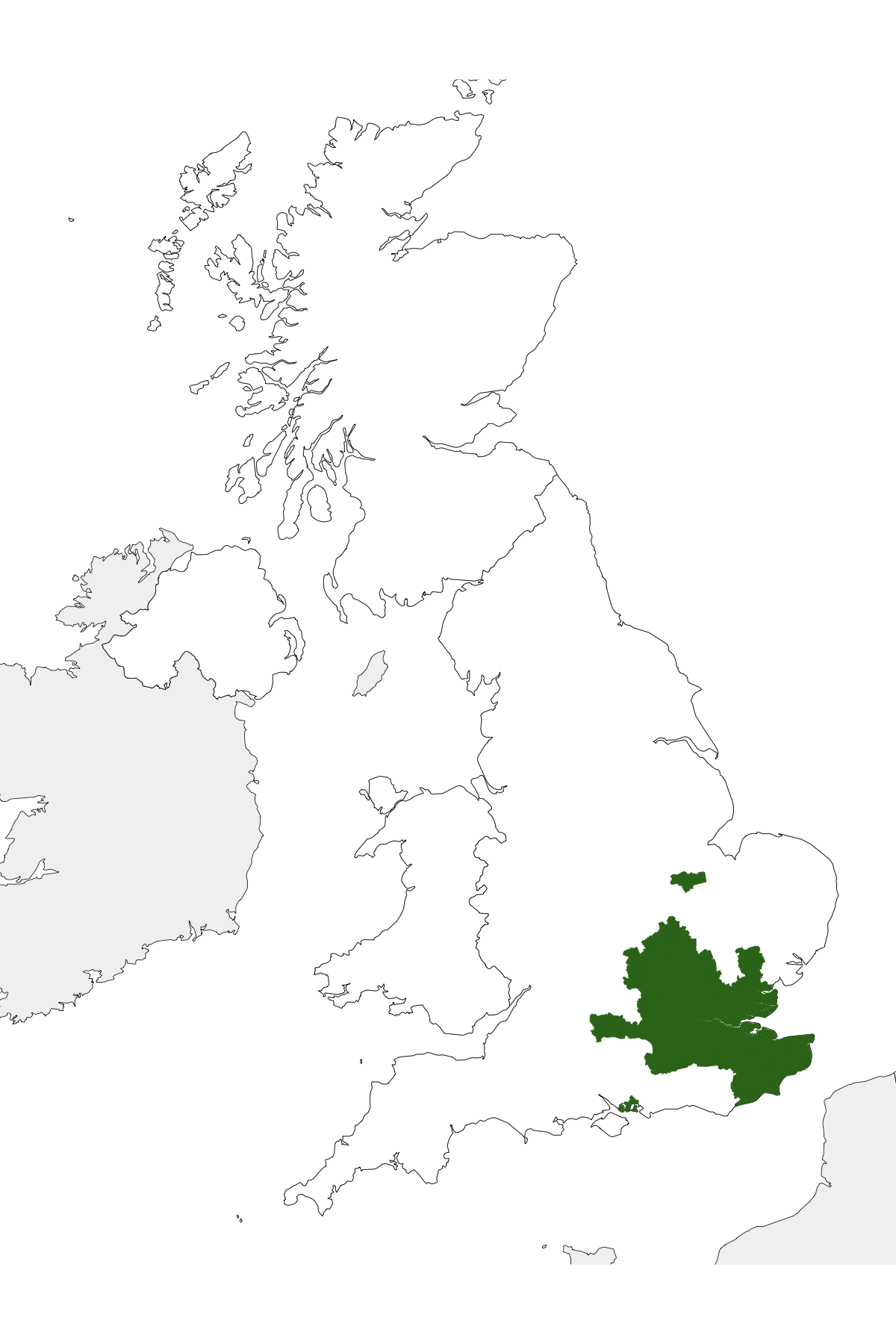

**Supplemental Figure 6. The extent of Tier 4 restrictions.** Restrictions limiting travel during the Christmas period were introduced on December 19th, 2020 and included a short term relaxation of measures on Christmas Day.

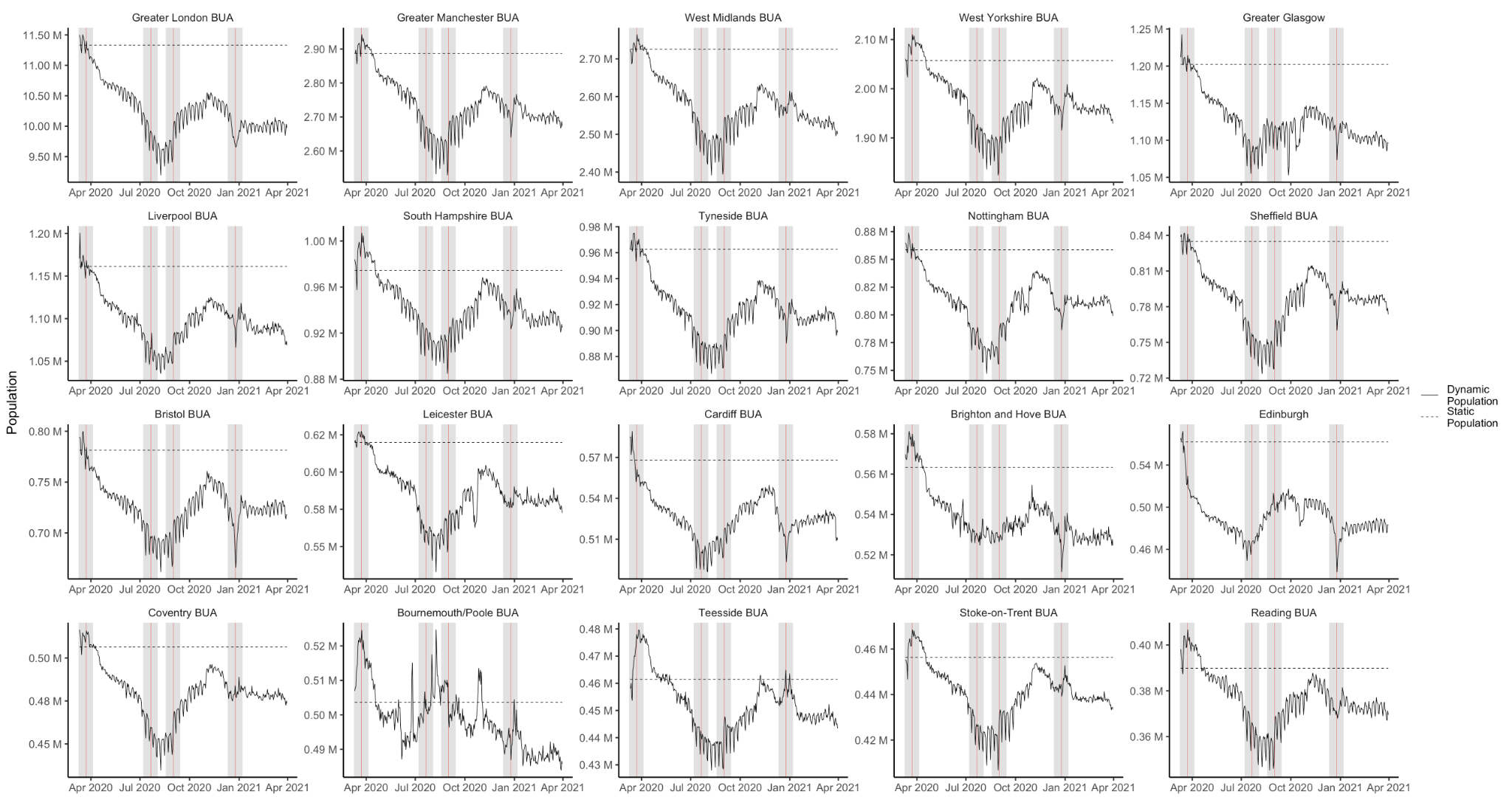

**Supplemental Figure 7. Time-varying population estimates for Built Up Areas.** Dynamic population estimates for the top 20 BUAs ranked by census population (from highest to lowest).

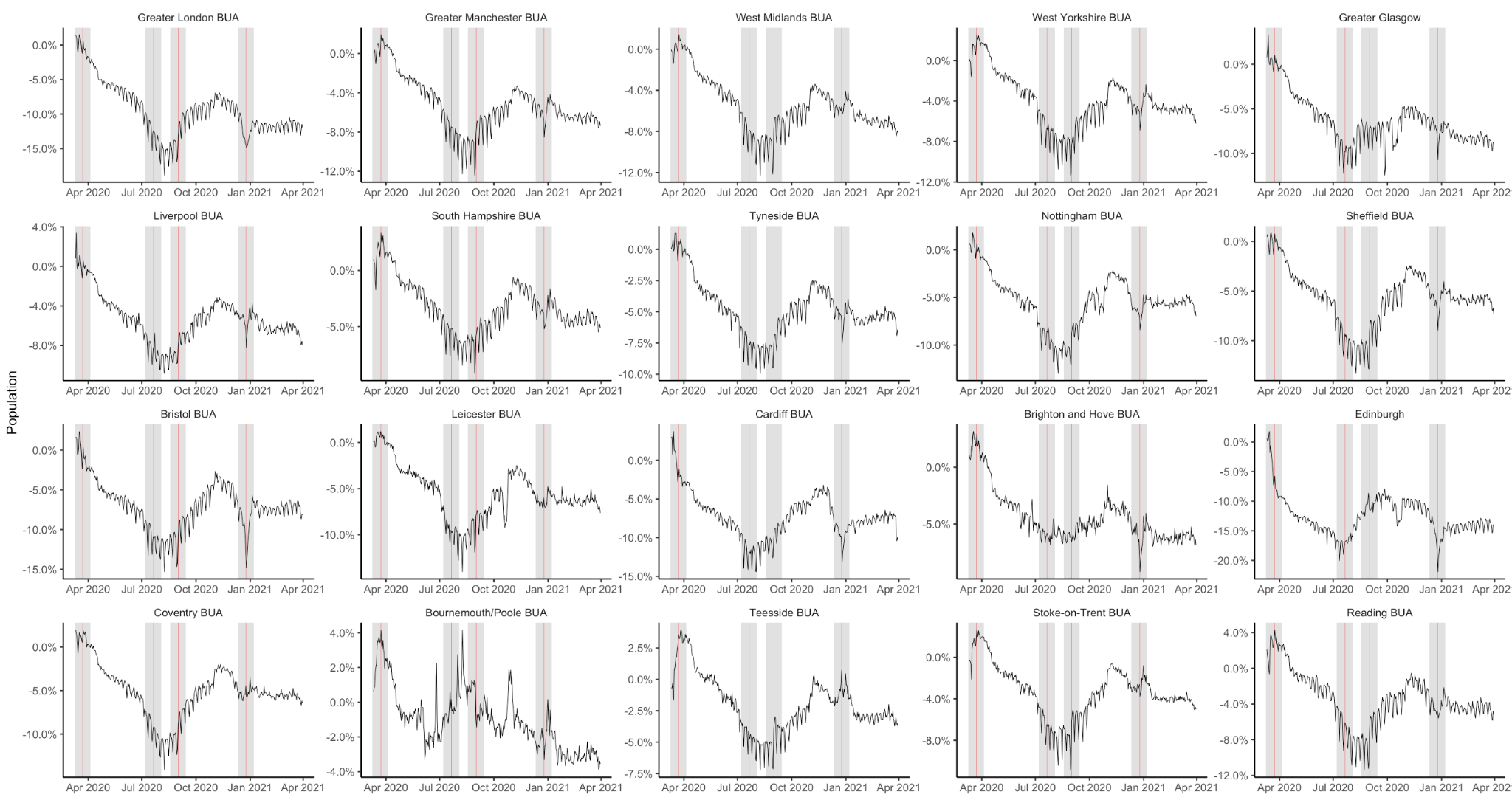

**Supplemental Figure 8. Time-varying percent population change estimates for Built Up Areas.** Dynamic population estimates for the top 20 BUAs ranked by census population (from highest to lowest) showing the percentage change from census population.

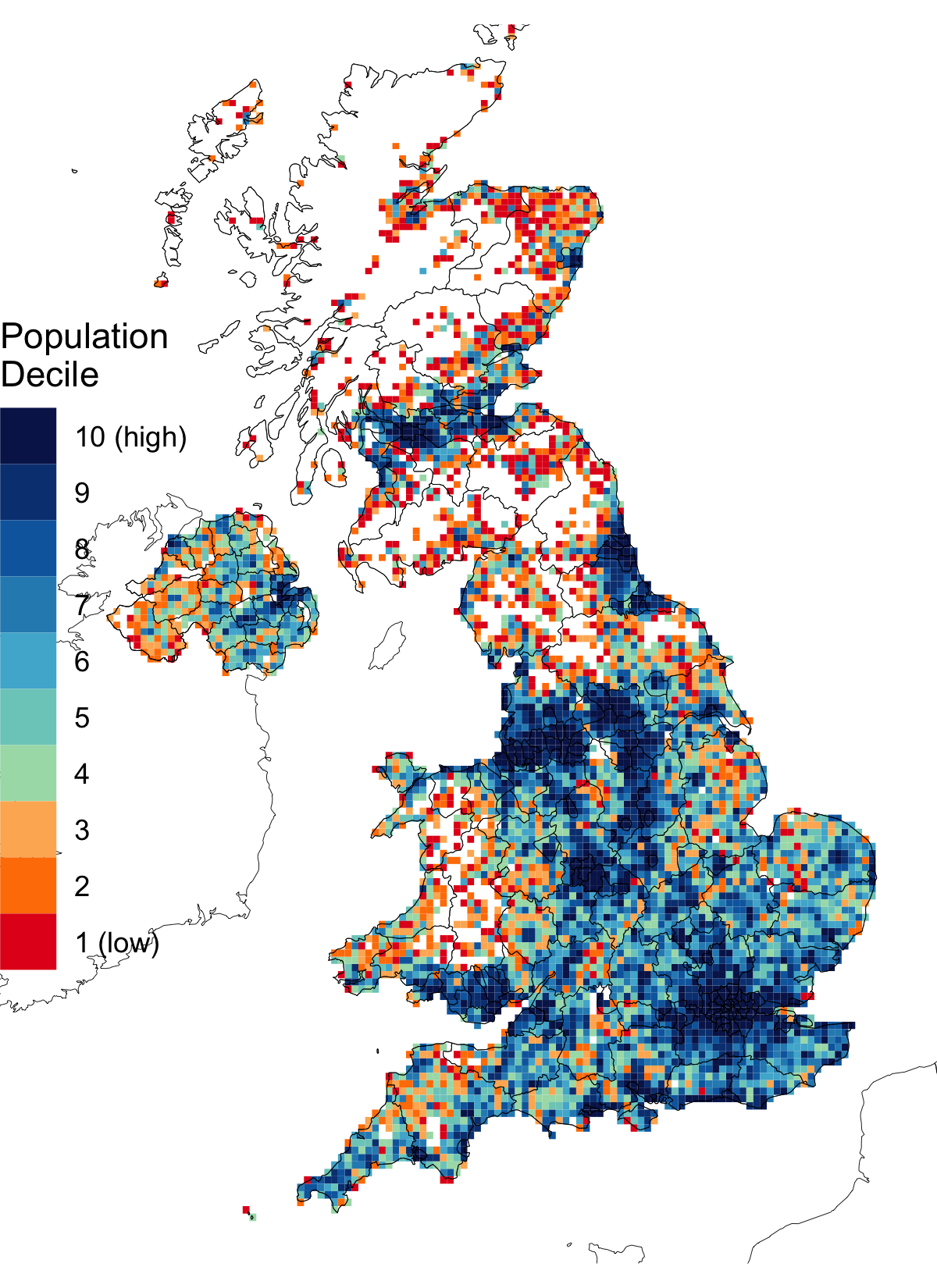

**Supplemental Figure 9. Tiles in population deciles.** The partition of tiles by population deciles based on the total population of each tile in the baseline period.

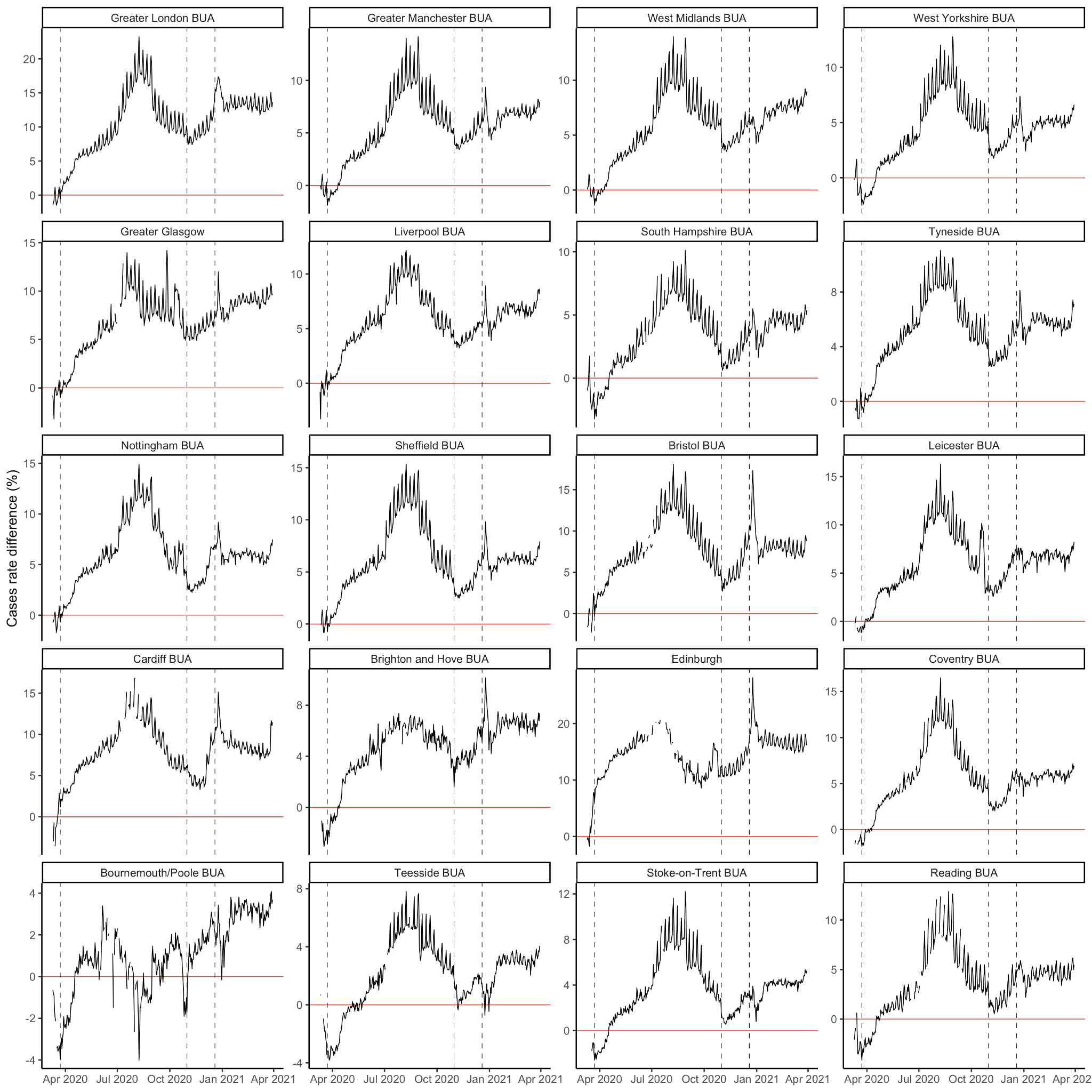

**Supplemental Figure 10. Differences in case rates for major BUAs.** The difference in rates of confirmed COVID-19 cases in the top 20 BUAs. Dates where the difference between rates is 0 show as gaps in the line.

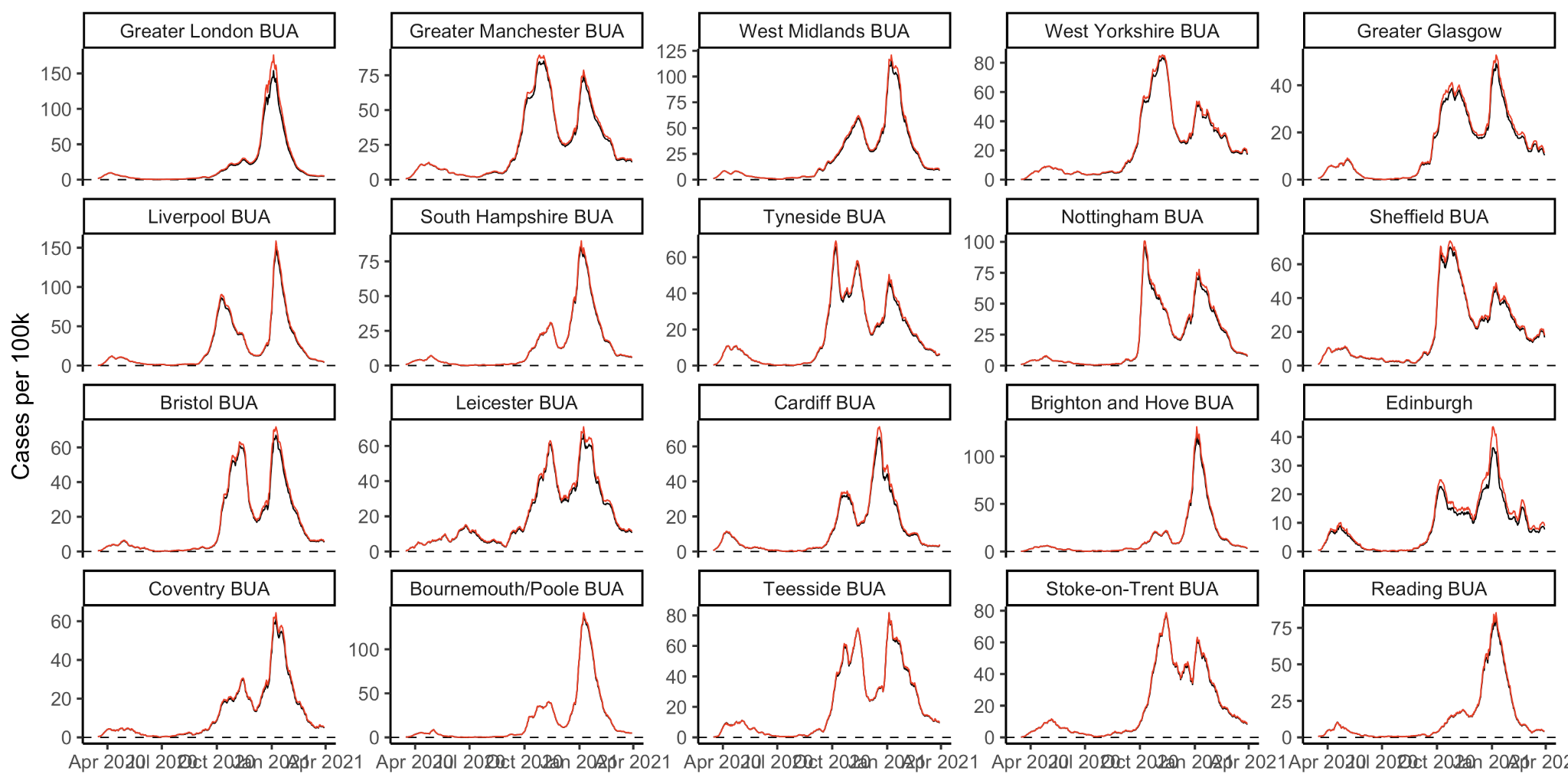

**Supplemental Figure 11. Rate of cases using static and dynamic populations.** The rate of confirmed COVID-19 cases in the 20 largest BUAs calculated using static populations and dynamic population estimates.
